# Association between Left Ventricular Hypertrophy and Neuroimaging Markers of Cerebral Small Vessel Disease in Patients with ischemic stroke: CNRS-III study

**DOI:** 10.1101/2023.02.08.23285679

**Authors:** Fangfang Yue, Hongyi Yan, Yuesong Pan, Jing Jing, Xia Meng, Yong Jiang, Zixiao Li, Hao Li, Yongjun Wang, Yilong Wang

## Abstract

Data from the CNSR III Study (The Third China National Stroke Registry), a total of 9600 participants were included in this study who underwent transthoracic echocardiography and brain magnetic resonance imaging. Our study demonstrated a strong association between left ventricular hypertrophy(LVH) and different neuroimaging markers of cerebral small vascular disease in patient with ischemic stroke. Concentric hypertrophy was associated with DWMH(deep white matter hyperintensity),EPVS (enlarged perivascular space),Lacune, brain atrophy, and CMBs(cerebral microbleeds), but not PVWMH(periventricular white matter hyperintensity). Eccentric hypertrophy was associated with DWMH,EPVS-CS(enlarged perivascular space in centrum semioval), Lacune, CMBs, but not PVWMH, EPVS-BG(enlarged perivascular space in basal ganglia), or brain atrophy.The results remained stable after adjustment for conventional covariates. There were some variance in two subtypes of LVH, especially in people with DWMH or EPVS-CS, eccentric LVH seems to have a higher risk than concentric LVH.

**Background:** Some clinical special cardiovascular diseases have a certain associationwith the occurrence and development of cerebral small vessel disease(CSVD). Left ventricular hypertrophy (LVH) as a subclinical heart damage preceded heart failure is considered to be a risk factor for CSVD ^[2.3]^. However, the precise relationship of LVH and CSVD is unclear.

**Methods:** Patients with acute ischaemic stroke or transient ischaemic attack in China from the registric survey of the CNSR-III (The Third China National Stroke Registry) from August 2015 to March 2018 were included in present study.The markers of CSVD on magnetic resonance imaging, including white matter hypertintensity, perivascular space, lacunes, cerebral microbleeds, and brain atrophy, were assessed at baseline survey.Participants were divided into 4 groups according to relative wall thickness and left ventricular mass index assesed by transthoracic echocardiography. The association of left ventricular geomatric patterns with CSVD imaging markers was analyzed using ordinary or binary logistic regression models.

**Results:** A total of 9600 participants were included in this study. The mean age of the participants was 61.9+11.2 years, and 6558(68.3%) were men.Compared with normal heart geometry, concentric hypertrophy had higher risk of deep white matter hyperintensity(DWMH)(odds ratio [OR]:1.54;95% confidence interval [CI]:1.16-2.04), enlarged perivascular space in basal gangalia(EPVS-BG) (OR:1.26; 95%CI:1.08-1.46), enlarged perivascular space in centrum semiovale(EPVS-CS) (OR:1.16; 95%CI:1.01-1.33), lacune (OR:1.45; 95%CI:1.26-1.67), brain atrophy (OR:1.42; 95%CI:1.20-1.69), but not periventricular white matter hyperintensity(PVWMH) after adjustment for covariates. Further, we did not found the association between eccentric heart hypertrophy and PVWMH, EPVS-BG and brain atrophy.

**Conclusions:** In this ischemic stroke registry,we found a possible association between LVH and CSVD imaging markers at different sites, neither concentric nor eccentric heart hypertrophy was associated with PVWMH, that was independent of hypertension and other traditional vascular risk factors.

**Graphical Abstract:** A graphic abstract is available for this article

## Introduction

Cardiovascular disease and cerebral small vessel disease (CSVD) are highly prevalent among elderly individuals, both leading to high mortality and morbidity and sharing many common vascular risk factors (VRFs) such as smoking, hypertension, and diabetes mellitus^[1]^. And also, some certain specific clinical cardiovascular disorders increase the risk of CSVD, such as coronary heart disease (CHD)^[4]^, atrial fibrillation (AF)^[5]^, and heart failure (HF)^[6]^. CVDs may contribute to CSVD by causing cerebral hypoperfusion, emboli, or infarcts^[1]^. However, the precise mechanism of CVDs and CSVD is unclearly. In addition, the neuroimaging markers differ, their underlying pathophysiological mechanisms may differ.

Chronic cerebral hypoperfusion is one of the attributable mechanisms for all neuroimaging markers of CSVD ^[7]^. Previous study indicated that left ventricular ejaction fraction(LVEF), a well-established cardiac perfusion indicator, is negatively correlated with the burden of CSVD in a dose–response manner^[8]^, but another study with 4366 samples found non-linear/u-shaped associations between LVEF and total brain and grey matter volume, showing that a higher LVEF is not necessarily better than lower LVEF, or a higher LVEF is not equal to higher brain perfusion. The same results were seen in studies examining the relationship between LVEF and cognitive function ^[8]^, which may be because a higher LVEF indicated left ventricular hypertrophy (LVH) --a pathological increase in left ventricular mass (LVM),whose stroke volume is lower^[9]^. Further,data from a large cohort ARIC (Atherosclerosis Risk in Communities) Study (5th ARIC visit) indicated that increased LVMI was associated with increased odds of brain infarction and more white matter hyperintensities^[10]^. Interestingly, in patients with heart failure with preserved ejection fraction (HFPEF), the left-ventricular wall is thickened ^[11]^. Therefore,we can known heart structure had changed before heart dysfunction. Above all, increased LVMI as a marker of atherosclerotic heart injury is more specific and sensitive in reflecting the connection of heart and brain than LVEF.

High blood pressure is the most consistent risk factor for all CSVD neuroimaging manifestations.^[12]^. Increased LVMI and LVH are clinical markers of hypertension-mediated heart damage^[32,33]^ and constitute surrogate indicators of the degree or duration of exposure to hypertension. ^[13,14]^. However, LVH is not present in all hypertensive patients, a review analysis has reported that the prevalence of LVH is between 36% and 41% in hypertensive populations this discrepancy may be due to genetic factors ^[34]^.One study by Razavi indicated that nearly one-fifth of the association of systolic BP to cognitive function is explained by LVMI^[15]^. Increased LVMI leaded to cognitive impairment mainly mediated by CSVD beyond clinical blood pressure^[16]^. Thus, LVH itself is probably the first step in the pathogenesis of hypertensive cardio-cerebral vascular disease.

Consequently, we investigated the association of LVH and different neuroimaging markers of CSVD in a relatively large-scale population registry (n=9600). We aim to provide new insights into the cardio-cerebral mechanisms and treatment of cerebral small vessel disease.

## METHODS

### Study Population

We utilized data from the CNSR-III registry(The Third China National Stroke Registry) which is a nationwide, multicenter, prospective registry of acute ischemic cerebrovascular events from 201 hospitals that cover 22 provinces and four municipalities in China between August 2015 and March 2018. Details about the population and enrollment of CNSR-III have been published previously^[17]^. A total of 9600 individuals who underwent transthoracic echocardiographic for examing LVH, brain MRI examination for CSVD image interpretation were included in this analysis. Subjects who had contraindications to magnetic resonance imaging (MRI), severe damage to the structure of the heart(eg: heart failure, dilated cardiomyopathy), severe cardiac hemodynamic changes(eg: atrial fibrillation, heart valve disease, thrombotic endocarditis, LVEF<50%) were excluded from the study.We collected baseline data on subject demographics, risk factors (drinking, and smoking status), medical history (diabetes, hypertension, dyslipidemia, heart disease, stroke, and TIA), admitted NIHSS score, and medication use (antihypertensive, lipid lowering, antidiabetic, antiplatelet, and anticoagulant). Baseline data were collected through face-to-face interviews by trained research coordinators. Hypertension was defined as either self-reported hypertension previously diagnosed by a physician or current use of antihypertensive agents or systolic blood pressure ⩾140 mmHg or diastolic blood pressure ⩾90 mm Hg. Diabetes was defined as a self-reported diabetes, pre-viously diagnosed by a physician or current use of antidiabetic agents or fasting plasma glucose ⩾7.0 mmol/L or 2-hour post-load glucose ⩾ 11.1 mmol/L or hemoglobin A1c ⩾6.5%.

### Measurement of LVH

The following protocols are recommended for all patients : transthoracic echocardiography,Left ventricular end-diastolic diameter (LVDD), Ventricular septal thickness(VST), and posterior wall thickness(PWT), LV ejection fraction(LVEF), Left atrium anterior and posterior diameter (LAD). Relative wall thick(RWT)was calculated according to this formula: 2×PWT/LVDD, and a value over 0.42 centimeters was defined as an increased RWT^[18].^ LV mass was calculated with a validated formula: LV mass =0.8(1.04[(VST+LVDD+PWT)3-LVDD3] +0.6)^[19]^, and indexed for body surface area (BSA). Mosteller’s formula is employed: BSA= (((Height in cm) x (Weight in kg))/3600)1/2). LVH was defined as LV mass index >115 g/m2 in men or >95 g/m2 in women[20].Respectively four categories of left ventricular geometry were defined: 1) normal (normal RWT and normal LVMI); 2) concentric remodeling (increased RWT and normal LVMI); 3) eccentric hypertrophy (normal RWT and increased LVMI); and 4) concentric hypertrophy (increased RWT and increased LVMI). Image data were collected in DICOM format on discs and analysed by the image research centre in Beijing Tiantan Hospital.

### Neuroimaging Makers of CSVD

The following brain magnetic resonance sequences are recommended for 9600 participants: T1 weighted, T2 weighted, Fluid-attenuated Inversion Recovery (FLAIR), Diffusion-Weighted Imaging (DWI) with Apparent Diffusion Coefficient (ADC) maps, SWI or T2* and Magnetic Resonance Angiography (MRA). CSVD neuroimaging markers include white matter hyperintensities,brain atrophy, lacunes of presumed vascular origin,moderately to severely enlarged perivascular spaces and microbleeds (only 5700 samples were available for CMBs interpretation owing to the lack of T2*/SWI) sequence). White matter hyperintensities in periventricular or deep brain were respectively graded by Fazekas scale (0 to 3) and brain atrophy were graded by GCA-scale (0 to 3)^[20]^. Lacunes were defined as asymptomatic 3- to 15-mm well-defined lesions that were located in the region of a perforating arteriole with the same signal characteristics as cerebrospinal fluid on T2- or T1-weighted MRI^[21]^. Enlarged perivascular space(EPVS) in CS or BG were defined as round, oval or linear-shaped lesions less than 3 mm in size with the same signal characteristics as cerebrospinal fluid and without a hyperintense rim ^[20]^. Moderately to severely EPVSs were categorized into a 10-20 group and >20 group by the number of EPVSs ^[22]^. CMBs were focal round lesions with low signal on T2-gradient echo or SWI images, with a size smaller than 10 mm ^[20]^. Image data were collected in DICOM format on discs and analyzed by neuroradiologists who were blinded to all clinical, echocardiographic information in Beijing Tiantan Hospital. The kappa coefficient of CSVD markers on brain MRI between raters were as follows: 82% for the presence of lacune, 88% for Fazekas scale of WMH, 89% for the severity of EPVS, and 85% for the presence of CMB.

### Statistical Analysis

Participants with the missing data of cystatin C and low MRI image quality were excluded. Categorical variables were presented as numbers and percentages, and were compared using the Chi-square test. Continuous variables were expressed as mean ± standard deviation, and were compared using t-test or one-way ANOVA. Binary logistic regression models were performed to evaluate the relationship of the patterns of left ventricular geometry with CSVD image markers. We estimated common odds ratio (OR) with their 95% CIs for total PVWMH, DWMH, EPVS-BG, EPVS-CS, Lacune, brain atrophy, and microbleeds.Two models were conducted for each outcome. In model a, age, sex were adjusted.In model b, age, gender, smoking, heavy drinking,coronary heart disease, previous antiplatelet agents, lipid-lowering drugs,hypoglycemic drugs,antihypertensive drugs use, LVEF and LAD were adjusted.Statistical analyses were performed using SAS 9.4 software (SAS Institute).

## RESULTS

### Patient Characteristics

Of 9600 patients were included in the present analysis. The mean age of the study subjects was 61.9 ± 11.2 years, and 6558(68.3%) were men. The flow chart of patient enrollment and exclusion criteria is in Figure 1. In our study of 9600 patients with normal LVEF, appropriately 49% (4737) of people were observed to have normal LV geometry, 31% with concentric remodeling (CR), 8% with eccentric left ventricular hypertrophy(EH), and 12% with concentric left ventricular hypertrophy(CH).The baseline characteristics of the participants are listed in Table 1. The participants with LVH tended to be older, be male, and had higher prevalence of hypertension, diabetes, current smoking, heart disease, higher proportion of medicine use,lower LVEF, and larger LAD than those with normal heart geometry.(Table 1). In addition, presence of PWHM, DWHM,moderately to severely EPVS-BG, moderately to severely EPVS-CS, brain atrophy, and lacunes respectively was 97.15%, 90.95%,31.31%, 48.06 %, 75.83%, and 50.38%. Of the 5771 participants who had SWI/T2* sequence, 1569(27.44%)had at least one CMBs. (Table 1).

**Table 1.**
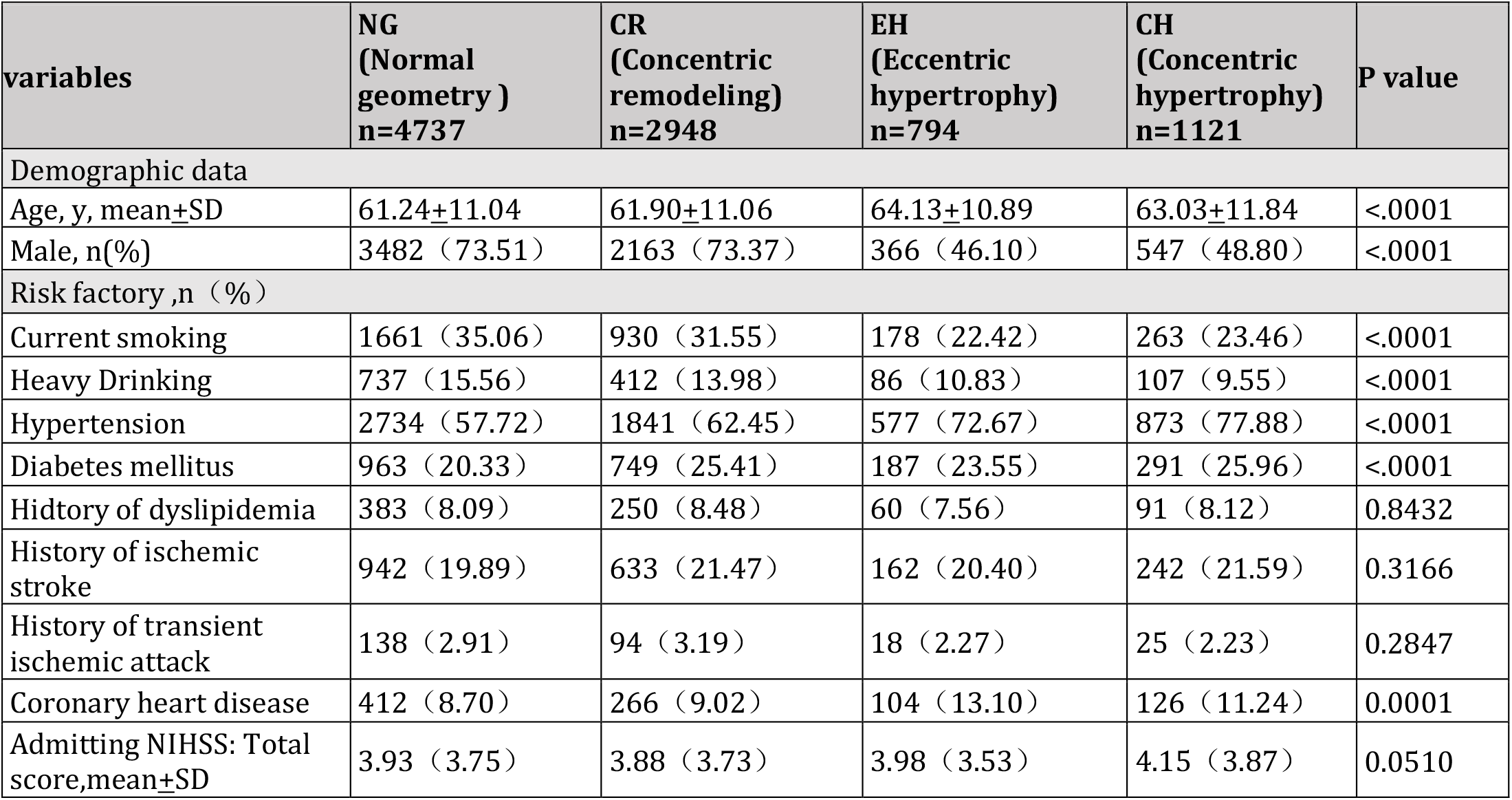
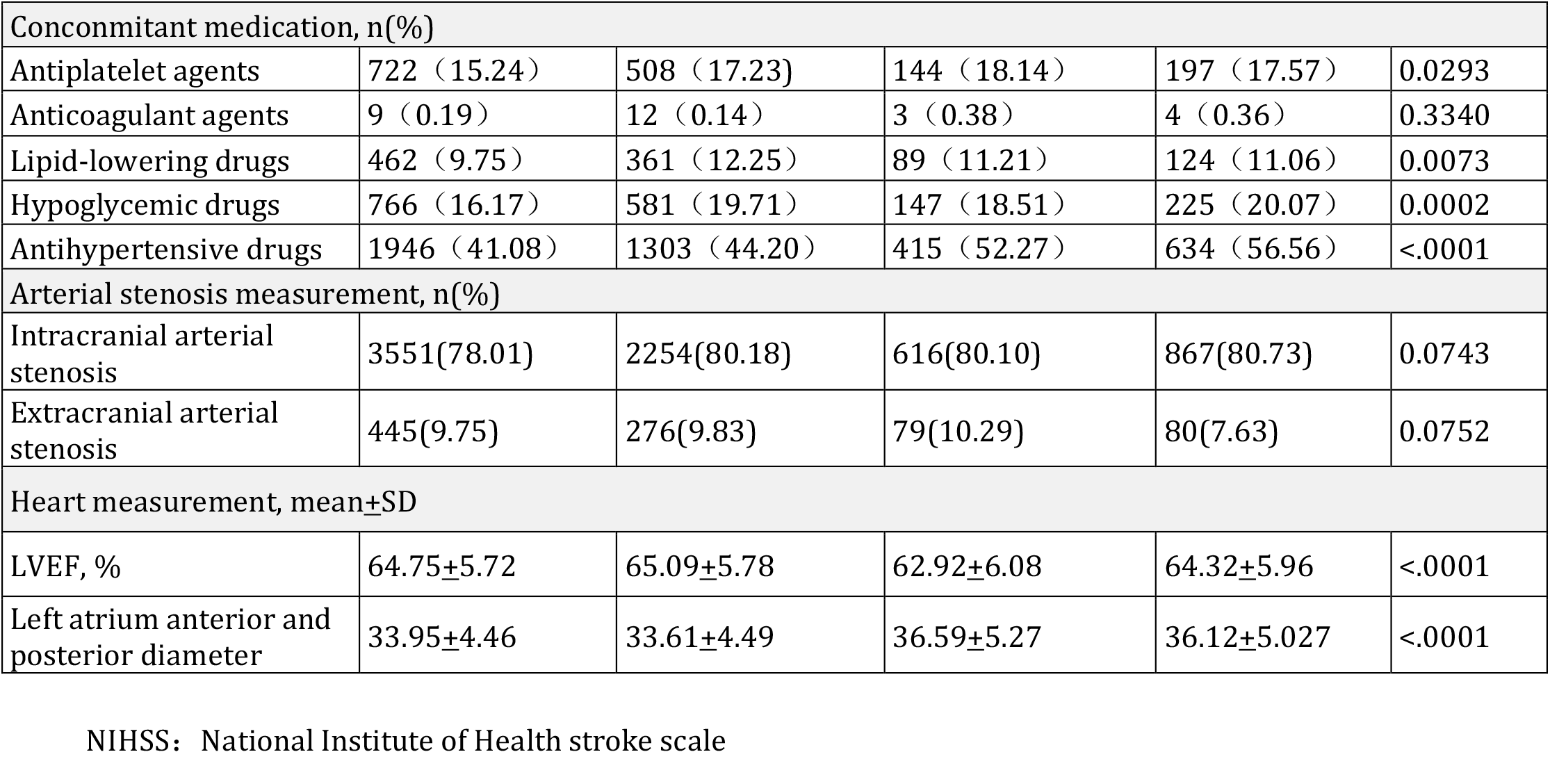
Demographic and Clinical Characteristics According to left ventricular geometry patterns

**Figure 1.**
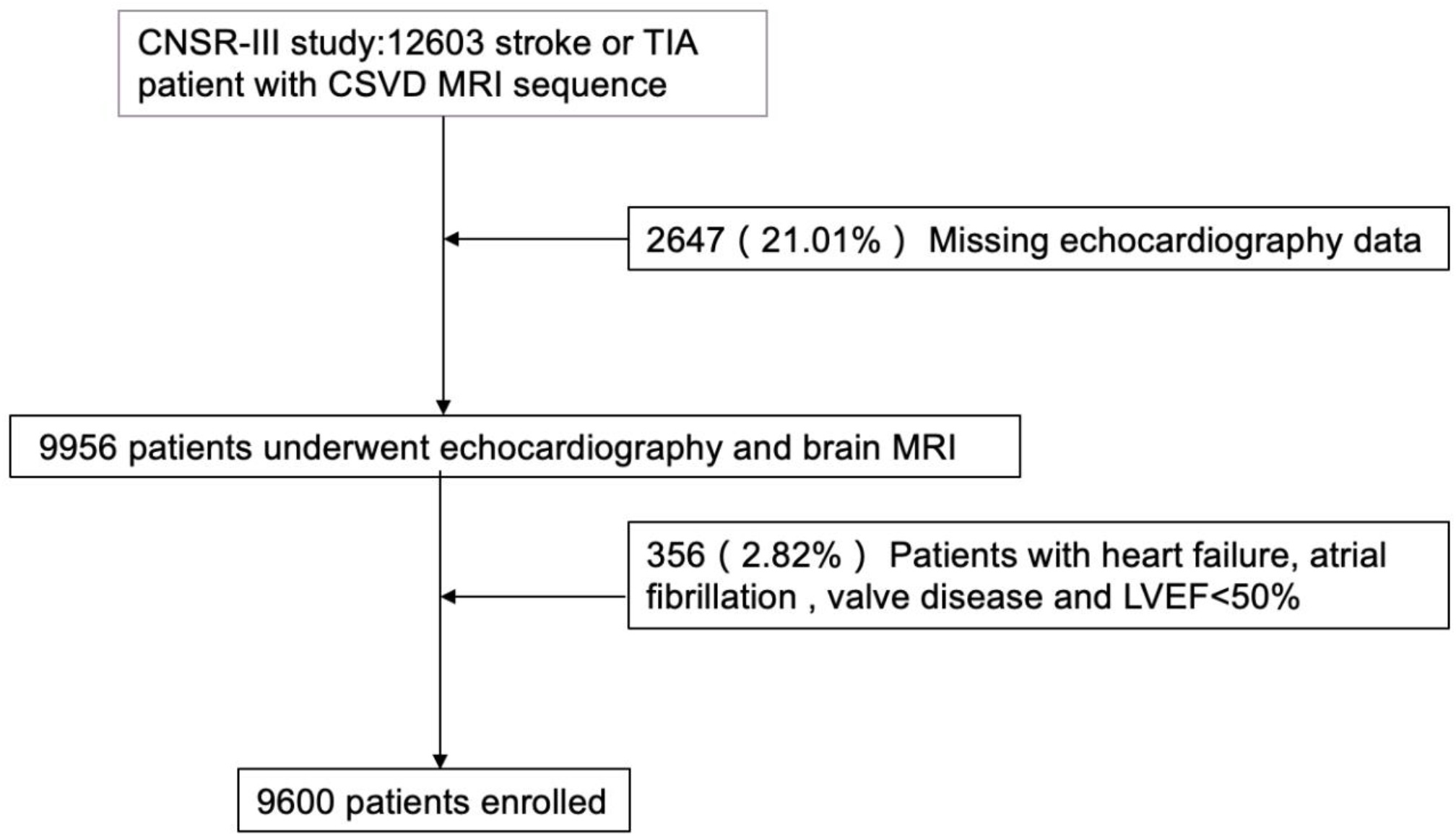

### LVH and WHM, EPVS location

The distribution of CSVD imaging markers by heart geometric patterns among all included participants is shown in table 2. Presence of PWHM was 96.83% in normal LV geometry, 96.71% in concentric remodeling, 98.87% in eccentric LVH, 98.39% in concentric LVH. Presence of DWMH was 89.61%, 90.91%, 94.58%, 94.11% respectively. After adjustment for age and sex (model a), LVH, both with eccentric(P=0.0009, OR:1.75;95%CI:1.26-2.44) or concentric(P<0.0001,OR:1.79;95% CI:1.36-2.35) pattern was associated with DWMH and both with eccentric(P=0.0170, OR:2.30;95%CI:1.16-4.56) and concentric(P=0.0184, OR:183;95% CI:1.11-3.01)hypertrophy was associated with PWMH. But after adjustment for pertinent covariates(p<0.05), LVH was associated with DWMH (P=0.0026,OR:1.54;95% CI:1.16-2.04 in concentric hypertrophy. P=0.0057, OR:1.61;95%CI: 1.15-2.25 in eccentric hypertrophy) not with PWMH.

**Table 2.**
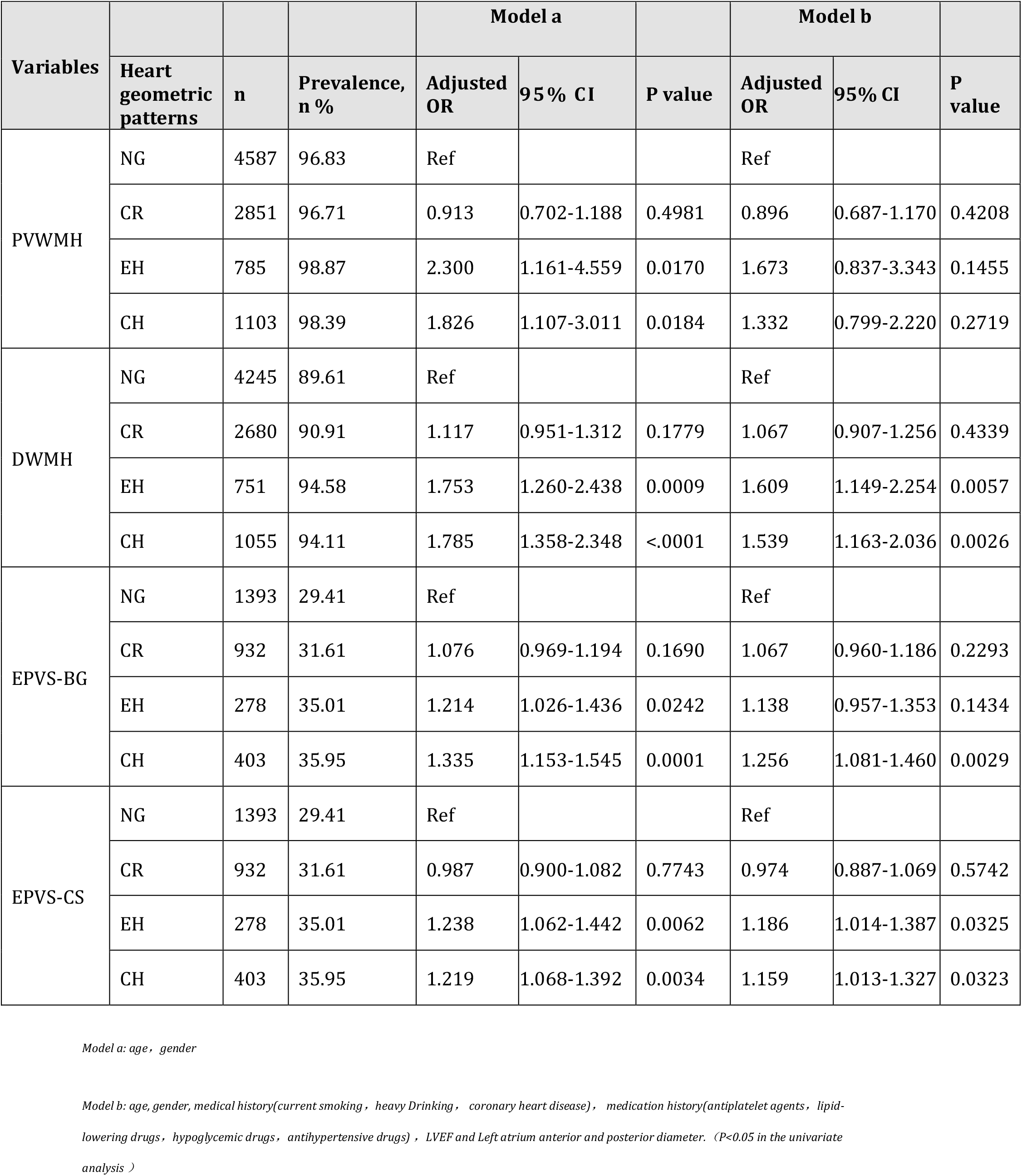
The Association of Heart Geometric Patterns and Withe Matter Hyperintensity and Enlarged Perivascular Space

**Table 3.**
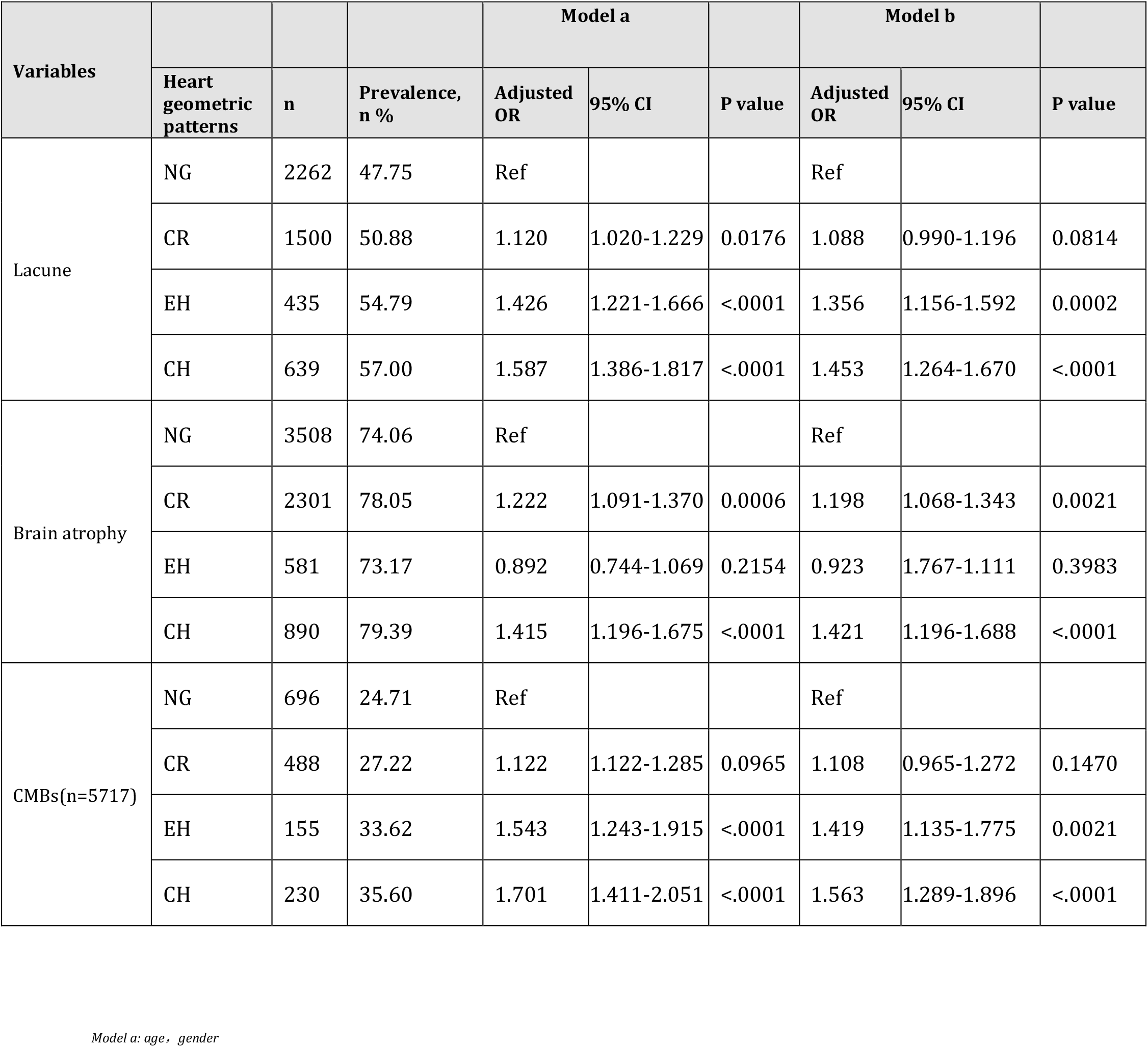
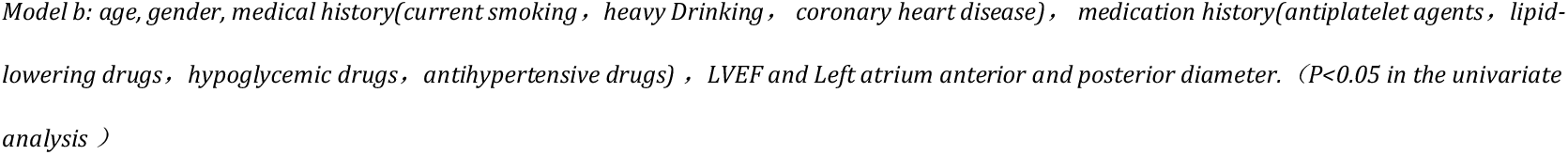
The Association of Heart Geometric Patterns and Lacune,brain atrophy, and CMBs.

Presence of moderate and severe EPVS-BG was 31.31%(3006). There were 1393(29.41%) participants with moderate and severe EPVS-BG in normal LV geometry, 932(31.61%) in concentric remodeling, 278(35.01 %) in eccentric LVH, 1103(98.39%) in concentric LVH. (Table 2).There were 2240(47.29%), 1388(47.08%), 411(51.76%) and 575(51.29%) participants with moderate and severe EPVS-CS respectively in four subgroups. After adjustment for conventional covariates, only concentric hypertrophy was associated with EPVS-BG(P=0.0029,OR:1.256;95%CI:1.081-1.460),while the association between both eccentric and concentric LVH and EPVS-CS was still observed, (P=0.032,OR:1.16;95% CI:1.01-1.33 in concentric hypertrophy. P=0.033, OR:1.19;95%CI:1.01-1.39 in eccentric hypertrophy)

### LVH and Lacune, brain atrophy, and CMBs

Concentric hypertrophy was associated with Lacune(OR:1.45;95% CI:1.26-1.67), brain atrophy(OR:1.20;95% CI:1.07-1.34), and CMBs(OR:1.56;95%CI:1.29-1.90), as compared with normal geometry of heart. Eccentric hypertrophy was associated with Lacue(OR:1.36;95%CI:1.16-1.60), CMBs(OR:1.42;95%CI:1.14-1.78)), but not brain atrophy.

## DISCUSSION

Our findings demonstrated a strong association between echocardiographically determined LVH and different CSVD neuroimaging markers but not PVWMH in ischemic stroke population. The results remained stable after adjustments for age, sex, hypertension, LEVF, LAD, and other vascular risk factors. Similar to heart geometric distribution of our study(49% with normal LV geometry, 31% with CR, 8% with EH, and 12% with CH),the four heart geometric proportion of a large single institution study of 35,602 patients with normal LVEF was 54% with normal geometry, 35% with CR, 5% with EH, and 6% with CH.

About the association among LVH geometries and WMH locations, unlike previous studies that concentric LVH has a greater detrimental consequence than eccentric LVH, our study found that eccentric LVH may carry higher risk for DWMH than concentric LVH. Several pathways may account for this connection. First, they have different vascular architecture, DWMH lesions are primarily supplied by long microvessels which are affected more likely by small vessel occlusion, while PWMH are supplied by short penetrating microvessels which are affected more directly by hypertension and traditional risk factors associated with stroke. ^[23]^. This makes it clear that our study did not see a association between LVH and PWMH after adjusting for hypertension and traditional risk factors. Secondly, the traditional paradigm was that concentric LVH was a response to increased after load(pressure overload), and that eccentric LVH was mainly the result of increased preload states (volume overload)^[24,25,26]^,which is easily understanded the result eccentric LVH carried higher risk than concentric LVH in DWMH. This provides new ideas and evidence for the mechanism exploration of WMH.What’s more, similar to the result of WMH location, eccentric LVH seemed to have higher risk than concentric LVH for participants with EPVS-CS. These findings are novel but do not conflict with the current understanding of PVS, a lot of studies suggested anatomical structures of PVS are different between the basal ganglia and the centrum semiovale^[27,28]^, Shigeki Yamada etal reported that EPVS-CS was not winding or branching like the BG-PVS ^[29]^. Different mechanisms may account for EPVS according to their anatomical distribution. In one study, 7-T MRI-visible PVS correlate spatially with arterioles^[28]^.lenticulostriate artery passing through the basal ganglia has a lager flow volume and highter pulse pressure wave amplitude than the superficial arteriole passing through the brain surface. Therefore, the impact of arterial pulsation, which is the main driving force of the fluid flow in the PVS^[30]^, might be larger in the basal ganglia rather than that in the centrum semiovale. Thus, EPVS-BG was proposed as an imaging biomarker associated with hypertensive arteriopathy or arteriosclerosis ^[31]^. This is account for our study result, the association between LVH and EPVS-BG was only found in concentric LVH rather than eccentric LVH beyond history of hypertension and antihypertension mediation use. LVH may itself be an independent risk factor for CSVD. We hypothesis that LVH may contribute CSVD by reducing cardiac functional efficiency. When the left ventricle hypertrophies, the electrical tracts in the myocardium lengthen, increasing the risk for cardiac arrhythmias. As the synchronized conduction of the heart becomes compromised, risk for blood stasis increases, potentially leading to thrombus formation^[35]^. A 2014 meta-analysis of 27,141 patients in 10 studies showed an 11.1% risk of supraventricular tachycardia (including AF) in those with LVH vs. 1.1% risk in those without (p<0.001)^[36]^. In a study from Japan, LVH-AF link was found to be strongest in those with eccentric and concentric LVH^[37]^. Multicenter Swiss-AF cohort study has demonstrated participants with atrial fibrillation had a substantial burden of CSVD detected on brain MRI:30%, 22% and 99% had evidence of either large noncortical or cortical infarcts or small noncortical infarcts, microbleeds and white matter lesions, respectively ^[38]^.

Moreover, an RCT study of 7020 patients with type 2 diabetes(median observation time, 3.1 years) found that the patients received empagliflozin, as compared with placebo, had a lower rate of the cardiovascular event(38% relative risk reduction), stroke and of death from any cause(32% relative risk reduction)^[39]^.And one of the underlying mechanisms behind the observed benefits possibly involve ameliorating adverse LV remodeling^[40]^.LVH Improving cardiac function may also have a brain protective effect.

## Strengths and limitations

Our study was the first to demonstrate the relationship between heart geometric patterns and location of WMH and EPVS. However, there were some limitations in our study. Firstly, stroke patients are thought of as having higher vascular risk factors burden than general people. Thus, the relationship between LVH and CSVD may be more obvious in these patients, although they have not been studied yet. Secondly, lack of image data analysis of lacune location or image sequence of CMBs, we only analyzed 5717 samples to exploring their relationship, and we didn’t analyze the association between LVH and CSVD total burden.

## CONCLUSION

Our study demonstrated the relationship between heart geometric patterns and neuroimaging markers of CSVD. There were some variance in two subtypes of LVH, especially in people with DWMH or EPVS-CS, eccentric LVH seems to have a higher risk than concentric LVH. More follow-up studies are needed to learn more underlying relationship of heart structure and neuroimaging markers of CSVD.

## Data Availability

The authors confirm that the data supporting the findings of this study are available within the article [and/or] its supplementary materials.

## Supplemental Material

STROBE Statement Checklist Figures 1

Tables 1-3

